# Genetic Evidence Causally Linking Pancreas Fat to Pancreatic Cancer: A Mendelian Randomization Study

**DOI:** 10.1101/2023.04.20.23288770

**Authors:** Hajime Yamazaki, Samantha A. Streicher, Lang Wu, Shunichi Fukuhara, Róbert Wagner, Martin Heni, Steven R. Grossman, Heinz-Josef Lenz, Veronica Wendy Setiawan, Loic Le Marchand, Brian Z. Huang

## Abstract

**Background & Aims:** Pancreatic ductal adenocarcinoma (PDAC) is highly lethal, and any clues to understanding its elusive etiology could lead to breakthroughs in prevention, early detection, or treatment. Observational studies have shown a relationship between pancreas fat accumulation and PDAC, but the causality of this link is unclear. We therefore investigated whether pancreas fat is causally associated with PDAC using two-sample Mendelian randomization.

**Methods:** We leveraged eight genetic variants associated with pancreas fat (P<5×10^-8^) from a genome-wide association study (GWAS) in the UK Biobank (25,617 individuals), and assessed their association with PDAC in the Pancreatic Cancer Cohort Consortium I-III and the Pancreatic Cancer Case-Control Consortium dataset (8,275 PDAC cases and 6,723 non-cases). Causality was assessed using the inverse-variance weighted method. Although none of these genetic variants were associated with body mass index (BMI) at genome-wide significance, we further conducted a sensitivity analysis excluding genetic variants with a nominal BMI association in GWAS summary statistics from the UK Biobank and the Genetic Investigation of Anthropometric Traits consortium dataset (806,834 individuals).

**Results:** Genetically determined higher levels of pancreas fat using the eight genetic variants was associated with increased risk of PDAC. For one standard deviation increase in pancreas fat levels (i.e., 7.9% increase in pancreas fat fraction), the odds ratio of PDAC was 2.46 (95%CI:1.38-4.40, P=0.002). Similar results were obtained after excluding genetic variants nominally linked to BMI (odds ratio:3.79, 95%CI:1.66-8.65, P=0.002).

**Conclusions:** This study provides genetic evidence for a causal role of pancreas fat in the pathogenesis of PDAC. Thus, reducing pancreas fat could lower the risk of PDAC.

## Introduction

With increasing body weight, fat is not only stored in the classical subcutaneous and visceral depots, but also accumulates within various organs ^1^. Excessive fat storage in the liver is a well-established risk factor for liver cancer, presumably through pro-inflammatory and pro-fibrotic mechanisms ^2^. While fat accumulation in the pancreas is a long-known phenomenon ^3^, it has received less attention due to past challenges in accurately quantifying pancreas fat in this small and irregularly shaped organ ^4^. Histologically, most fat in the pancreas is stored in adipocytes that reside between pancreas cells ^4^. Adipocytes in the pancreas secrete a variety of proteins, including chemokines and cytokines, thereby promoting tissue inflammation ^5^.

Recent developments in imaging methods such as magnetic resonance imaging (MRI) have not only revealed that pancreas fat accumulation is common ^6, 7^, but also enabled a better understanding of the clinical significance of pancreas fat ^8–10^. Several imaging studies have shown that individuals with higher levels of pancreas fat have an increased diabetes risk, presumably due to signals from local adipocytes impairing insulin secretion ^11, 12^. Most importantly, higher levels of pancreas fat are also hypothesized to cause pancreatic ductal adenocarcinoma (PDAC) ^4, 13^, which accounts for 90% of all pancreatic cancers and is expected to become the second-leading cause of cancer-related mortality by 2030 ^14^.

Evidence from a meta-analysis of cross-sectional studies and a recent case-control study with an observational period of 1-36 months have shown that pancreas fat is associated with precancerous lesions and PDAC ^15, 16^. However, due to the long latency of tumorigenesis, findings from observational studies may be subject to reverse causation bias. In this case, pancreas fat accumulation could be a consequence, rather than a risk factor of PDAC. For instance, it has been reported that in early-stage PDAC, fat accumulates only in close proximity to the tumor, but not in other parts of the pancreas ^17^.

Evaluating causal relationships can be accomplished using Mendelian randomization, a method that uses genetic variants (e.g. single-nucleotide polymorphisms [SNPs]) to assess the causal effect of an exposure (e.g., pancreas fat) on a disease (e.g., PDAC) ^18^. Mendelian randomization studies are considered natural randomized trials, in which the random inheritance of genetic variants works as random treatment assignments. Because genetic variants are assigned randomly at conception and are not affected by acquired diseases or environmental factors, Mendelian randomization is less prone to reverse causation and confounding ^19, 20^. We made use of this methodology and performed a Mendelian randomization study to evaluate the causal association of genetically determined pancreas fat with PDAC. We thereby aimed to clarify if pancreas fat is indeed a causal contributor to PDAC, which could ultimately lead to improvement in prevention, early detection, or treatment of this highly lethal cancer where only 10% of patients are alive in 5 years from diagnosis ^14^.

## Methods

### Study design and Source of data

We conducted a two-sample Mendelian randomization study using data from two large-scale genome-wide association studies (GWAS) for pancreas fat and PDAC ^21-23^. The details of assumptions required in Mendelian randomization are shown in **Figure 1**. All studies had been approved by ethical review boards and informed consent was obtained. **Table 1** shows descriptive information of the GWAS datasets used in this Mendelian randomization study.

**Figure 1.**
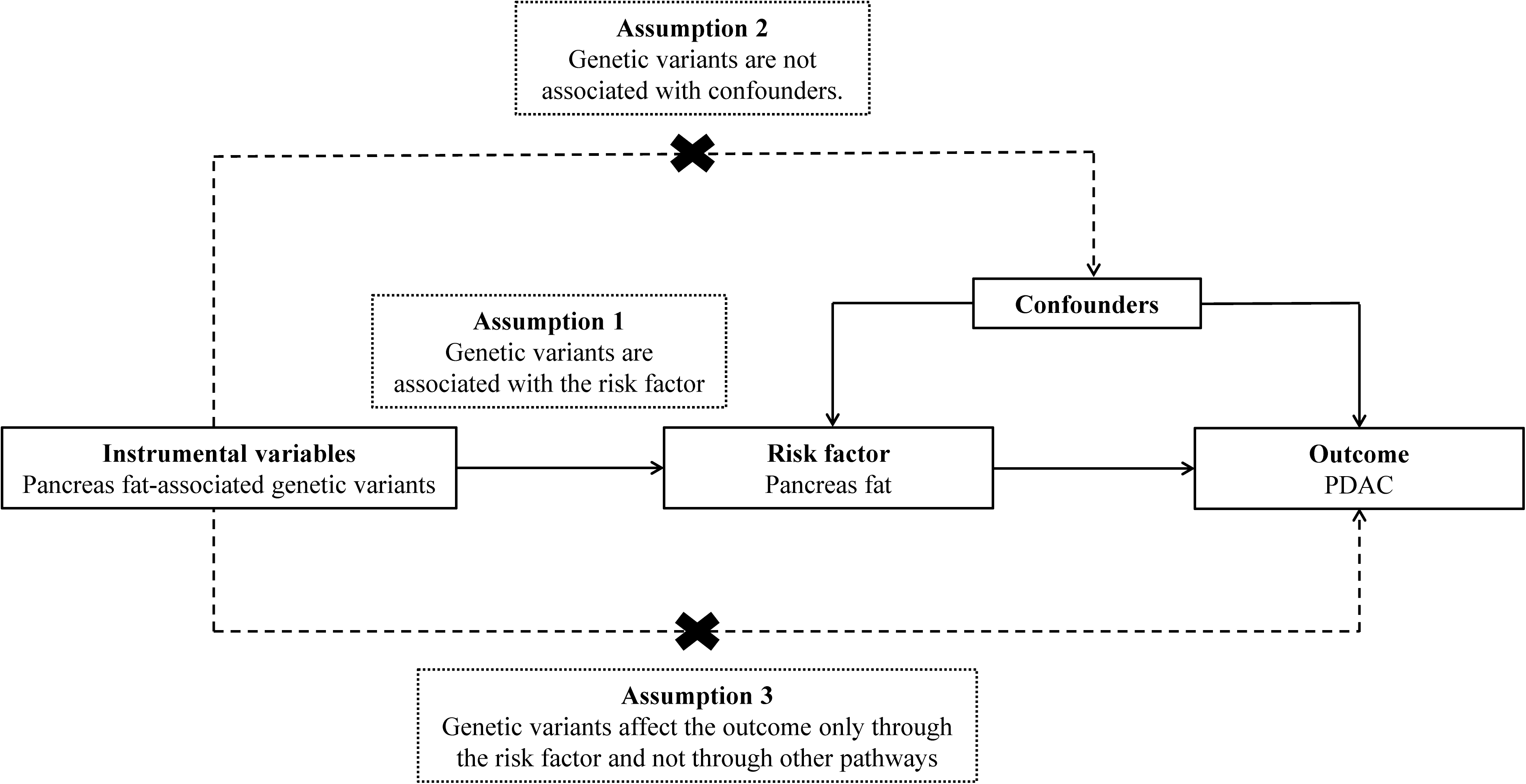
Schematic diagram illustrating the assumptions of Mendelian randomization analysis. The dashed lines represent violations of the Mendelian randomization assumptions. Abbreviations: PDAC, pancreatic ductal adenocarcinoma

**Table1.**
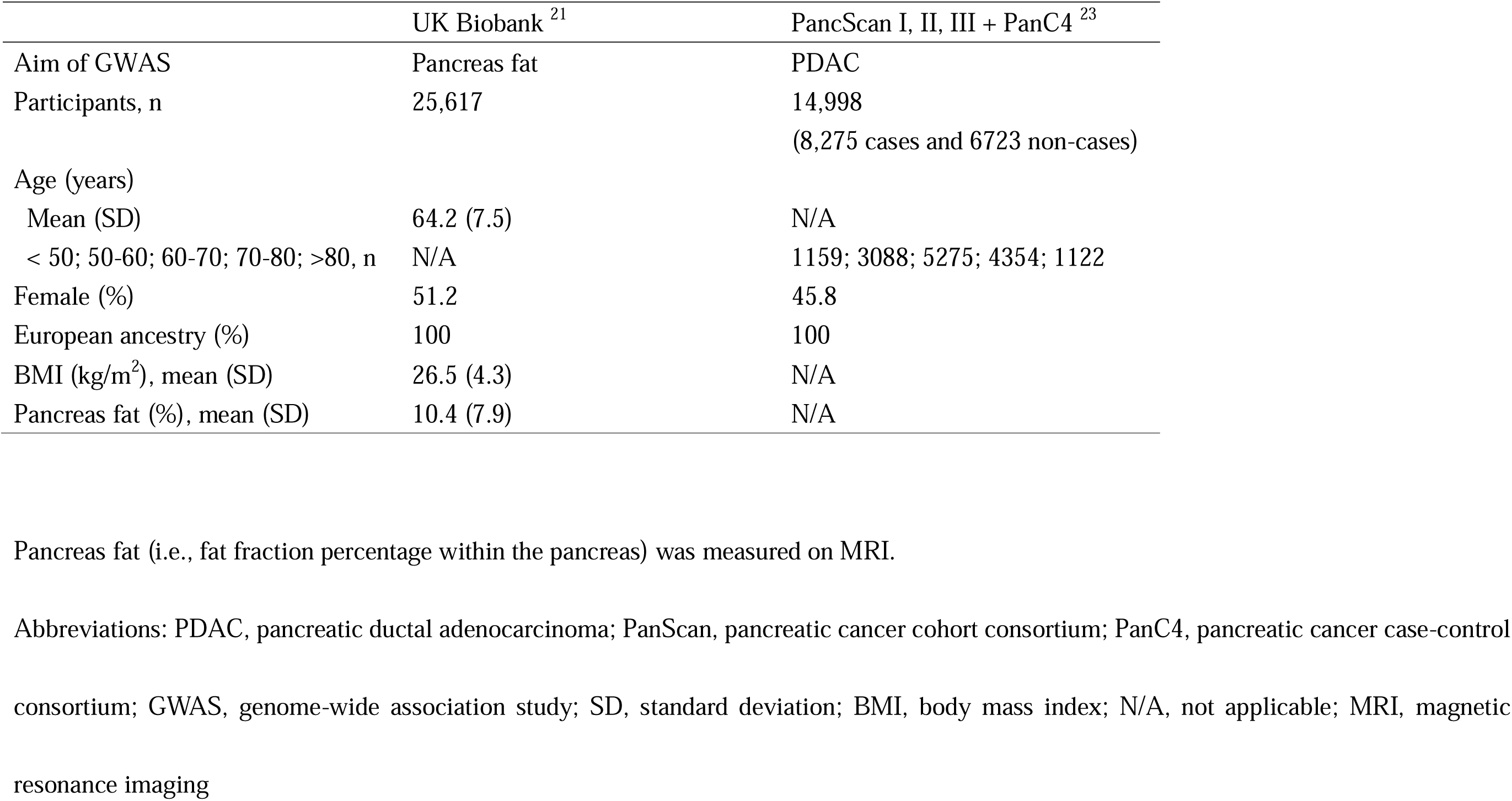
Descriptive information of genome-wide association studies used in this Mendelian randomization study

We obtained pancreas fat-related GWAS summary statistics from the UK Biobank (N = 25,617 individuals) ^21, 24^. In the GWAS study, pancreas fat levels were measured on MRI and shown as fat fraction percentage within the pancreas ^21^. Pancreas fat fraction measured on MRI represents histological pancreas fat fraction, defined as the percentage of pancreatic intraparenchymal fat in the total pancreatic parenchyma ^10^.

For PDAC genetic associations, we used information from the Pancreatic Cancer Cohort Consortium (PanScan) I, II, III and the Pancreatic Cancer Case-Control Consortium (PanC4) GWAS dataset (N = 14,998 individuals) ^23^.

As a sensitivity analysis to address potential pleotropic associations with body mass index (BMI), we used BMI-related GWAS summary statistics from a meta-analysis of the UK Biobank and the Genetic Investigation of Anthropometric Traits (GIANT) consortium (N = 806,834 individuals)^25^.

### Selection of genetic instruments

Genetic variants used in this Mendelian randomization study were selected as follows (**Figure 2**). All nine independent genetic variants associated with pancreas fat levels at genome-wide significance (P < 5.0 × 10^-8^) were selected from the UK Biobank GWAS, in which age, age squared, sex, imaging center, scan date, scan time, genotyping batch, and genetic relatedness were controlled for in the analysis ^21^. We further excluded one genetic variant (rs13040225) with a palindromic SNP (i.e., those where the alleles are complementary, G/C or A/T) and MAF above 0.4 to avoid ambiguity of effect direction. The remaining eight genetic variants were used for the main analysis of this Mendelian randomization study. Among the eight genetic variants, four were not found in the GWAS summary statistics for PDAC in the PanScanI-III/PanC4 GWAS. Therefore, we used the same proxy SNPs in linkage disequilibrium (r^2^> 0.7) for these genetic variants, as done in a previous Mendelian randomization study for pancreas fat and diabetes mellitus ^24^ (**Table 2**).

**Figure 2.**
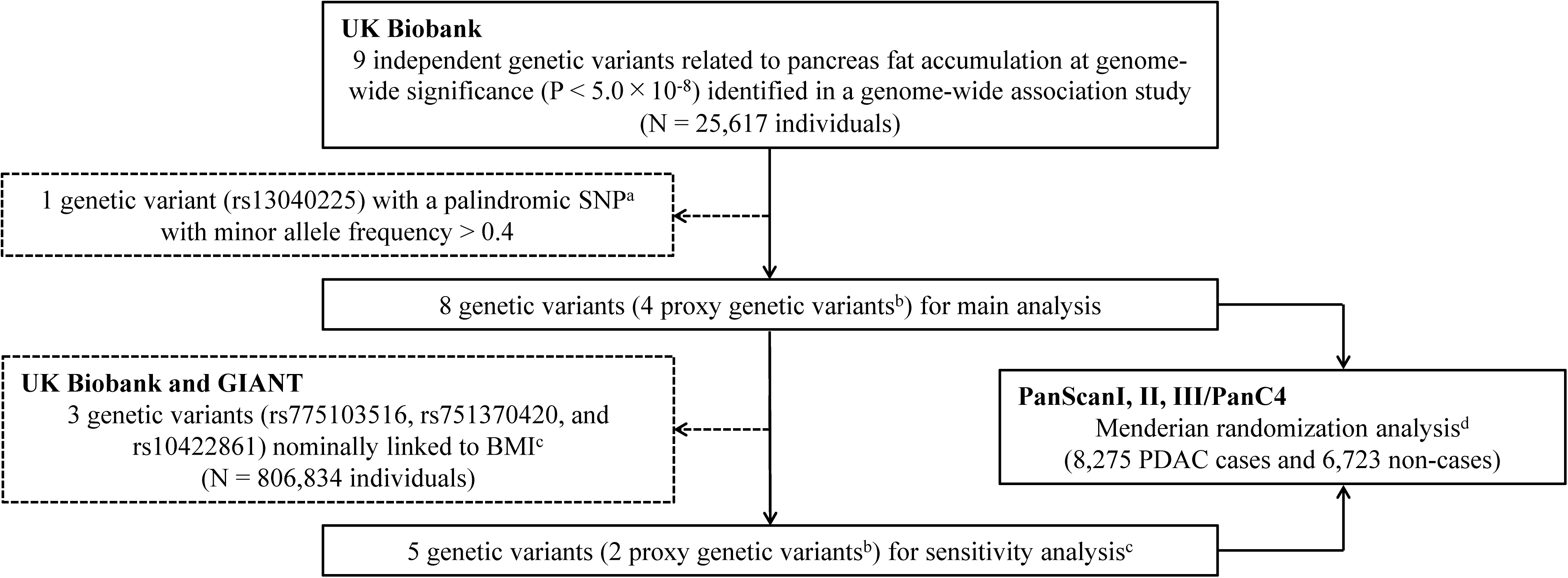
Data sources and selection of genetic instruments using Mendelian randomization. ^a^Palindromic SNPs are those where the alleles are complementary (G/C or A/T). ^b^Proxy genetic variants were used when selected genetic variants did not exist in PanScanI, II, III, or PanC4. ^c^Association of each genetic variant with BMI was evaluated using summary statistics obtained from meta-analysis results of the UK Biobank and GIANT consortium. Although none of the eight genetic variants were associated with BMI at the genome-wide significance, we further conducted a sensitivity analysis excluding the three genetic variants with a nominal BMI association. ^d^The estimates for the association of each genetic variant with PDAC were combined using the inverse-variance weighted method, with summary statistics for PDAC obtained from PanScan I, II, III, and PanC4. Abbreviations: SNP, single-nucleotide polymorphism; MAF, minor allele frequency; GIANT, genetic investigation of anthropometric traits; BMI, body mass index; PanScan, pancreatic cancer cohort consortium; PanC4, pancreatic cancer case-control consortium; PDAC, pancreatic ductal adenocarcinoma

**Table2.**
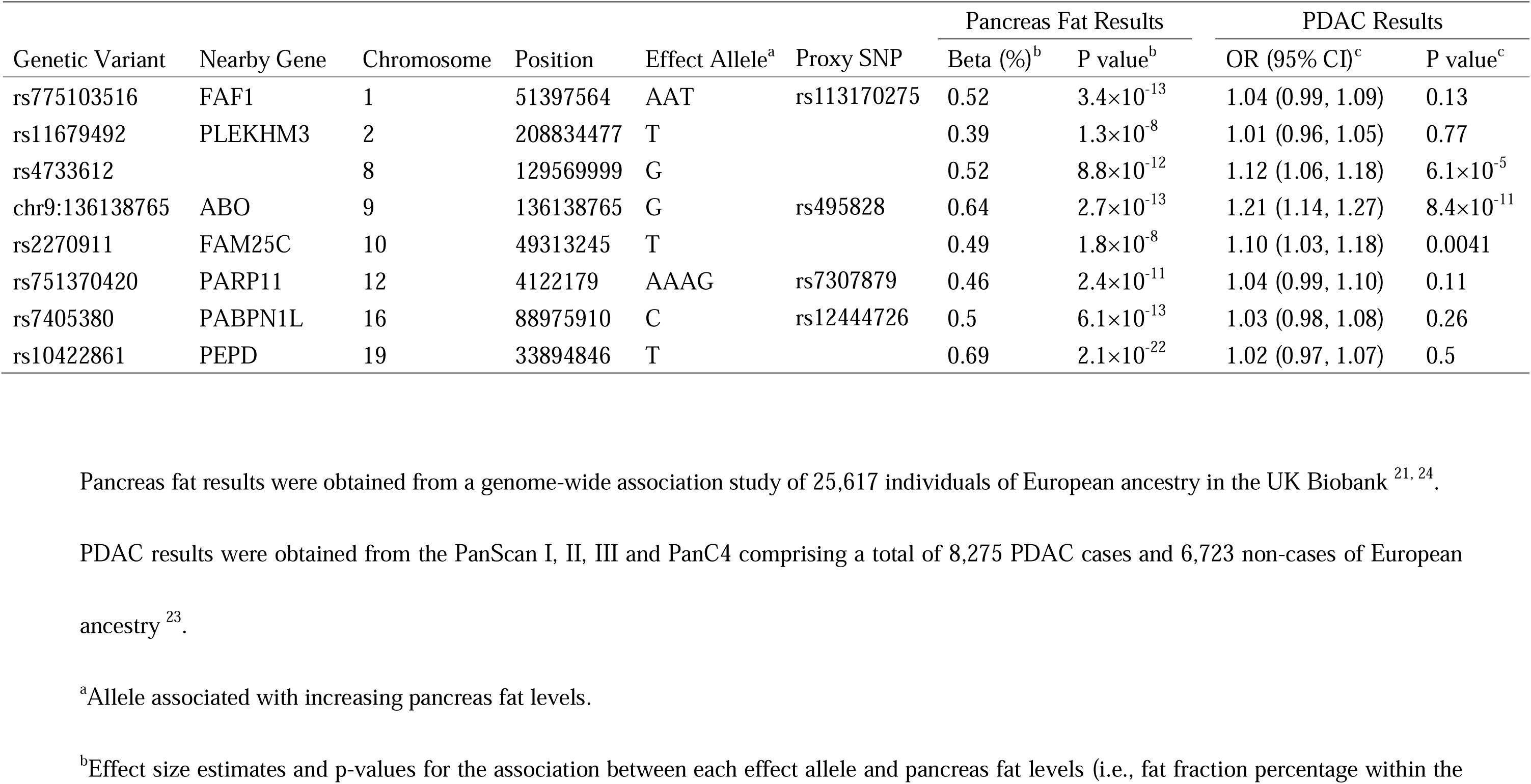
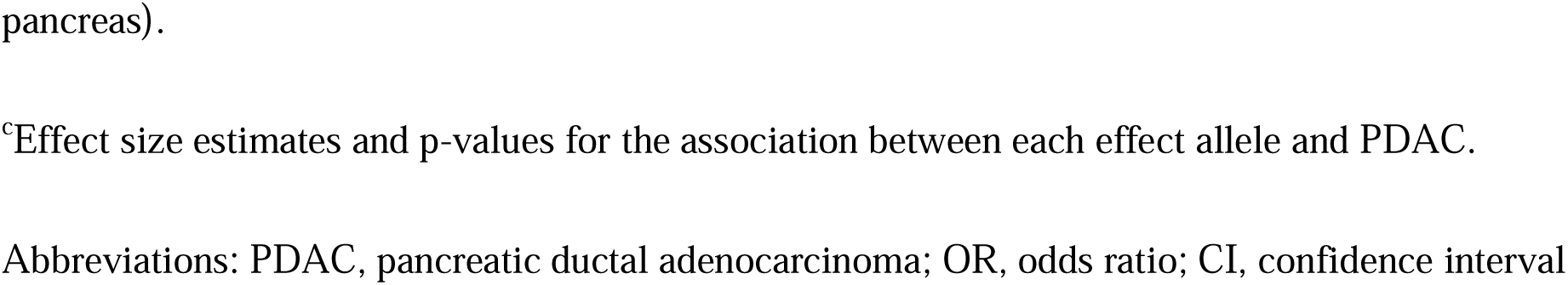
Characteristics of the pancreas fat-associated genetic variants

| Genetic Variant | Nearby Gene | Chromosome | Position | Effect Allele<sup>a</sup> | Proxy SNP | Pancreas Fat Results |  | PDAC Results |  |
| --- | --- | --- | --- | --- | --- | --- | --- | --- | --- |
|  |  |  |  |  |  | Beta (%)<sup>b</sup> | P value<sup>b</sup> | OR (95% CI)<sup>c</sup> | P value<sup>c</sup> |
| rs775103516 | FAF1 | 1 | 51397564 | AAT | rs113170275 | 0.52 | 3.4×10<sup>-13</sup> | 1.04 (0.99, 1.09) | 0.13 |
| rs11679492 | PLEKHM3 | 2 | 208834477 | T |  | 0.39 | 1.3×10<sup>-8</sup> | 1.01 (0.96, 1.05) | 0.77 |
| rs4733612 |  | 8 | 129569999 | G |  | 0.52 | 8.8×10<sup>-12</sup> | 1.12 (1.06, 1.18) | 6.1×10<sup>-5</sup> |
| chr9:136138765 | ABO | 9 | 136138765 | G | rs495828 | 0.64 | 2.7×10<sup>-13</sup> | 1.21 (1.14, 1.27) | 8.4×10<sup>-11</sup> |
| rs2270911 | FAM25C | 10 | 49313245 | T |  | 0.49 | 1.8×10<sup>-8</sup> | 1.10 (1.03, 1.18) | 0.0041 |
| rs751370420 | PARP11 | 12 | 4122179 | AAAG | rs7307879 | 0.46 | 2.4×10<sup>-11</sup> | 1.04 (0.99, 1.10) | 0.11 |
| rs7405380 | PABPN1L | 16 | 88975910 | C | rs12444726 | 0.5 | 6.1×10<sup>-13</sup> | 1.03 (0.98, 1.08) | 0.26 |
| rs10422861 | PEPD | 19 | 33894846 | T |  | 0.69 | 2.1×10<sup>-22</sup> | 1.02 (0.97, 1.07) | 0.5 |

Pancreas fat results were obtained from a genome-wide association study of 25,617 individuals of European ancestry in the UK Biobank <sup>21, 24</sup>.
PDAC results were obtained from the PanScan I, II, III and PanC4 comprising a total of 8,275 PDAC cases and 6,723 non-cases of European ancestry <sup>23</sup>.
<sup>a</sup>Allele associated with increasing pancreas fat levels.
<sup>b</sup>Effect size estimates and p-values for the association between each effect allele and pancreas fat levels (i.e., fat fraction percentage within the
pancreas).
<sup>c</sup>Effect size estimates and p-values for the association between each effect allele and PDAC.
Abbreviations: PDAC, pancreatic ductal adenocarcinoma; OR, odds ratio; CI, confidence interval

Considering possible pleotropic effects of the genetic variants on potential confounders, we also evaluated the association of the eight genetic variants with BMI using GWAS summary statistics of the UK Biobank and the GIANT consortium meta-analysis ^25^. None of the eight genetic variants were associated with BMI at a genome-wide significance threshold (P < 5.0 × 10^-8^). However, using the Bonferroni corrected threshold of P < 0.00625 ([P < 0.05]/8 genetic variants) as done in a previous Mendelian randomization study by Larsson, Burgess, and Michaelsson (2017) ^26^, three of the genetic variants (rs775103516, rs751370420, and rs10422861) had a nominal association with BMI. Therefore, we conducted a sensitivity analysis excluding these genetic variants to minimize potential residual pleiotropy.

### GWAS data for PDAC

GWAS data for PDAC in PanScan I, PanScan II, PanScan III, and PanC4 were downloaded from dbGaP (study accession nos.: phs000206.v5.p3 and phs000648.v1.p1). The detailed information for these data has been described in previous publications ^22, 27–30^. In brief, genotyping was performed on the Illumina HumanHap550, 610-Quad, OmniExpress, and OmiExpressExome arrays, respectively. Standard QC was conducted according to the guidelines recommended by the consortia ^22, 23^. Study subjects who were related to each other, had missing information on age or sex, had gender discordance, had non-European ancestry based on genetic estimation, or had a low call rate (less than 94% and 98% in PanScan and PanC4, respectively) were excluded. Duplicated SNPs and those with a high missing call rate (of at least 6% and 2% in PanScan and PanC4, respectively), or violations of Hardy-Weinberg equilibrium (of P < 1×10^-7^ and P < 1×10^-4^ in PanScan and PanC4, respectively) were also excluded. In the PanC4 dataset, we also excluded SNPs that had a MAF < 0.005, more than one Mendelian error in HapMap control trios, or more than two discordant calls in study duplicates. SNPs with sex differences in allele frequency > 0.2 or in heterozygosity > 0.3 for autosomes/XY were further excluded. We conducted the genotype imputation with the Haplotype Reference Consortium reference panel (r1.1 2016), using Minimac4 after phasing with Eagle v2.4 ^31, 32^. Imputed SNPs with an imputation quality of > 0.3 were retained. All of the genetic variants used in this Mendelian randomization study had an imputation quality of > 0.8 except for rs2270911, which had an imputation quality of 0.6. The associations between individual SNPs and PDAC risk were further assessed with logistic regression adjusting for age, sex and the top 10 principal components. In the final analyses, we included 8,275 PDAC cases and 6,723 non-cases of European ancestry ^23^.

### Statistical analysis

To evaluate the strength of the association between each genetic variant and pancreas fat (assumption 1 in **Figure 1**), we calculated F-statistics. F-statistics should be more than 10 to be valid genetic variants for Mendelian randomization ^18^. Cochran’s Q value was calculated to evaluate the heterogeneity among estimates obtained using different genetic variants.

For the primary analysis, the association of each genetic variant with PDAC was weighted by its association with pancreas fat, and estimates were combined using the random-effects inverse-variance weighted (IVW) method. This method is the most efficient and provides valid causal estimates when the average pleiotropic effect is zero. If the genetic variants used in this study are additionally associated with another risk factor for PDAC (i.e., presence of pleiotropic effect), then either assumption 2 or 3 for Mendelian randomization in **Figure 1** is violated.

To account for potential pleiotropy, we conducted five sensitivity analyses in our Mendelian randomization analyses: IVW method with leave-one-out analysis, MR-Egger regression method, weighted median method, MR-PRESSO, and IVW method after exclusion of the three genetic variants nominally linked to BMI. For leave-one-out analysis, each genetic variant was excluded, and we used the IVW method on the remaining genetic variants to evaluate the causal association of genetically determined pancreas fat with PDAC. Under the Instrument Strength Independent of Direct Effect (InSIDE) assumption ^33^, MR-Egger can provide valid causal estimates even if the average pleiotropic effect is not zero, and the intercept in MR-Egger can be tested to judge whether pleiotropic effects exist. However, the drawback of MR-Egger is the wide confidence intervals (CI). Weighted median method can provide valid causal estimates even if up to 50% of genetic variants have pleiotropic effect. MR-PRESSO can detect outlier genetic variants and calculate IVW estimates after exclusion of the outliers ^19^. Lastly, to address potential pleiotropy with BMI, a potential confounder, we excluded the three genetic variants nominally linked to BMI and conducted IVW analysis.

We considered P < 0.05 as statistically significant for our Mendelian randomization analyses. Statistical analyses were performed using Stata 17 (StataCorp, College Station, TX) and R version 4.0.5 (R Foundation for Statistical Computing, Vienna, Austria). We used the MendelianRandomization package ^34, 35^ and MR-PRESSO package ^36^ in R to conduct the Mendelian randomization analysis.

## Results

Characteristics of the eight pancreas fat-associated genetic variants are shown in **Table 2**. All of the eight genetic variants were strongly associated with pancreas fat: mean F-statistics 54 (min 33, max 103). About 1.6% of the variation in pancreas fat levels was explained by the eight genetic variants. The odds ratios (OR) of PDAC for all eight genetic variants were greater than the reference value of one.

In the primary Mendelian randomization analysis, genetically determined pancreas fat levels were associated with PDAC risk (**Figure 3)**. The OR of PDAC per one standard deviation (SD) increase in genetically determined pancreas fat level (i.e., per 7.9% increase in pancreas fat fraction) was 2.46 (95% CI: 1.38, 4.40; P = 0.002), an average 146% increased risk of PDAC per one SD (7.9%) increase in pancreas fat.

**Figure 3.**
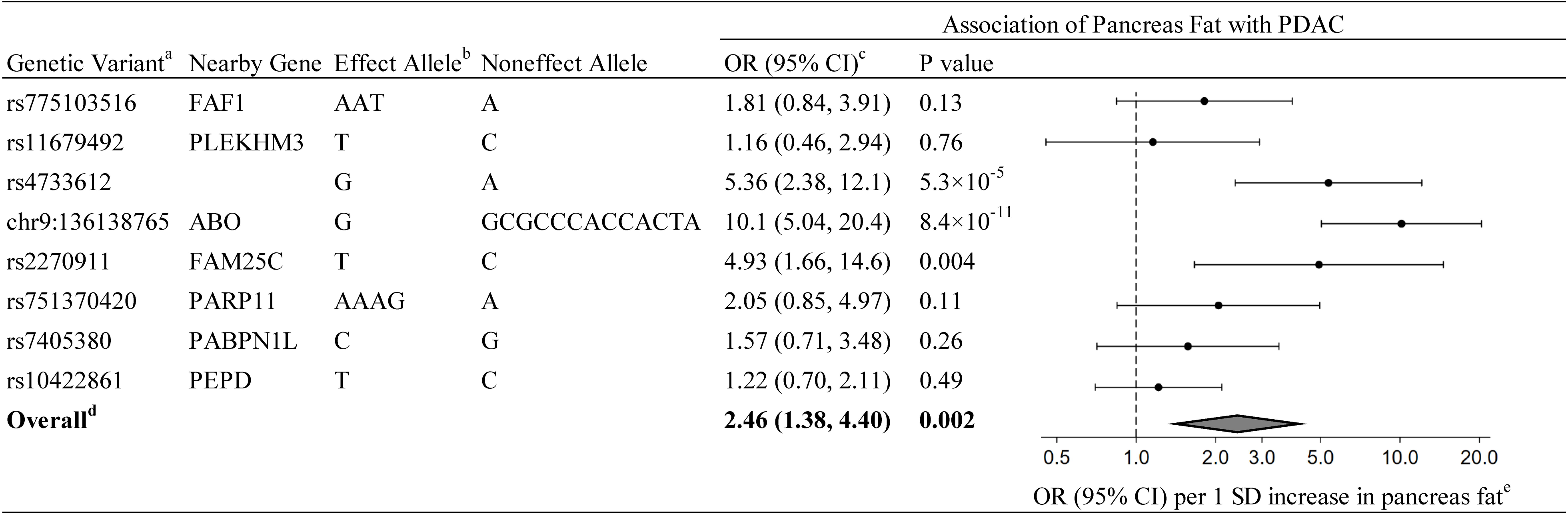
Primary Mendelian randomization estimates of the association between pancreas fat and PDAC. ^a^Proxy single-nucleotide polymorphisms were used for rs775103516 (rs113170275), chr9:136138765 (rs495828), rs751370420 (rs7307879), and rs7405380 (rs12444726). ^b^Allele associated with increasing pancreas fat levels. ^c^OR (95%CI) of PDAC per 1 SD increase in genetically determined pancreas fat levels (i.e., per 7.9% increase in pancreas fat fraction). ^d^Random-effects inverse-variance weighted method was used to obtain the overall estimate for the association of genetically determined pancreas fat with PDAC. ^e^Data markers indicate the OR for the association of genetically determined pancreas fat with PDAC, which was estimated using each genetic variant. Error bars indicate 95% CIs. Abbreviations: PDAC, pancreatic ductal adenocarcinoma; OR, odds ratio; CI, confidence interval; SD, standard deviation

The Mendelian randomization sensitivity analyses also showed consistent association between genetically determined pancreas fat levels and PDAC (**Table 3**). All of the leave-one-out ORs and 95% CIs indicated a statistically significant association with PDAC (**Supplementary Figure 1**). This result was the same even after excluding rs2270911 which had a relatively low imputation quality. The weighted median method and MR-PRESSO method showed similar associations (OR [95% CI]: 1.79 [1.13, 2.83] and 2.29 [1.61, 3.26], respectively) while the MR-Egger method showed a wide CI (OR 4.56 [95% CI: 0.14, 144.9]). Using MR-PRESSO, we found evidence of outliers (P_global test_ < 0.001), but the Mendelian randomization estimates for PDAC did not alter the inference of the results after removal of the outliers (P_distortion_ = 0.57). Although Cochrane’s Q value was high (31.6), there was no evidence of pleiotropy in the MR-Egger method (MR-Egger intercept, -0.042; P = 0.72). After removal of the three genetic variants nominally linked to BMI, the association between genetically determined pancreas fat levels and PDAC remained significant (OR [95% CI]: 3.79 [1.66, 8.65], P = 0.002).

**Table 3.**
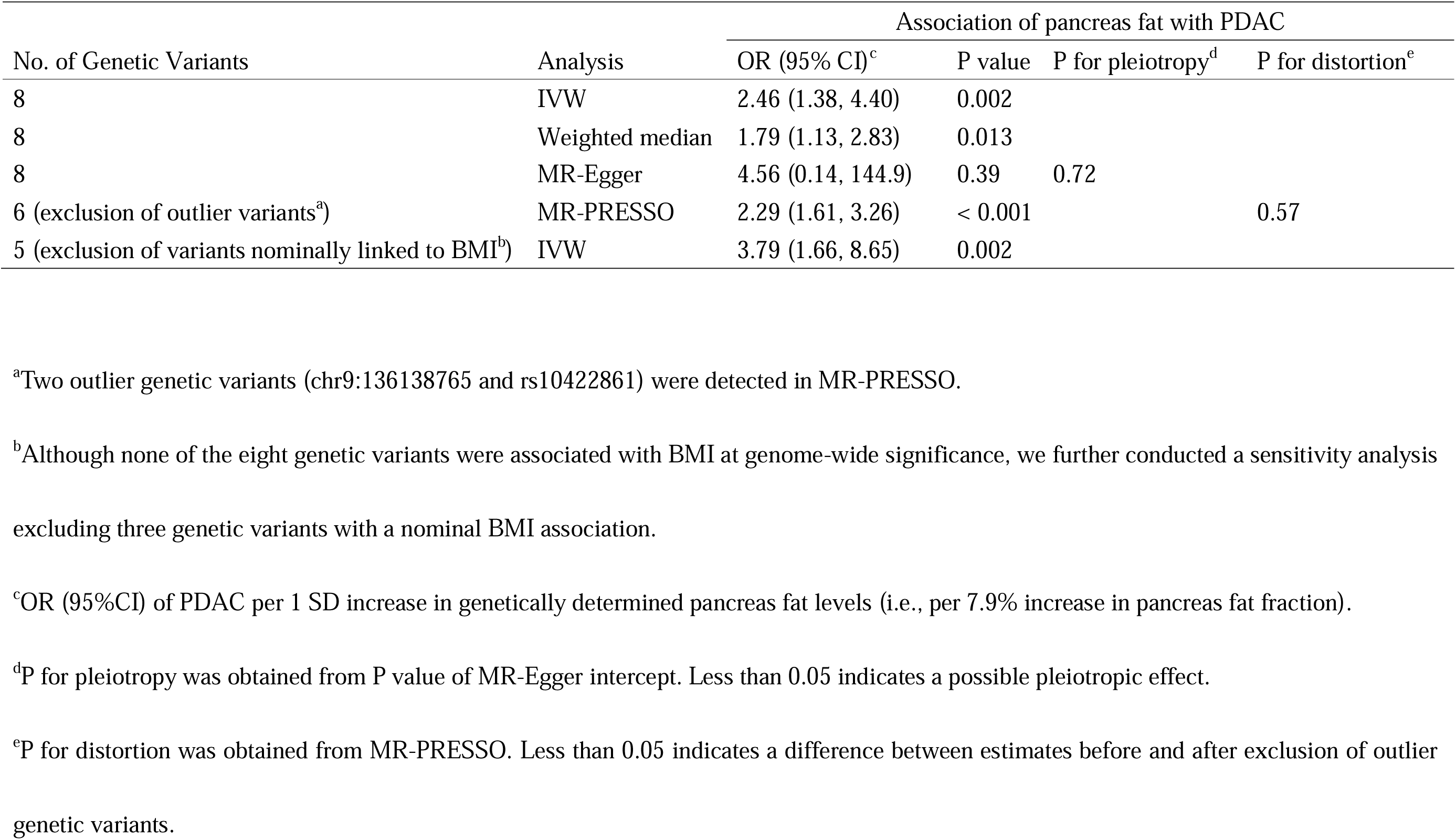
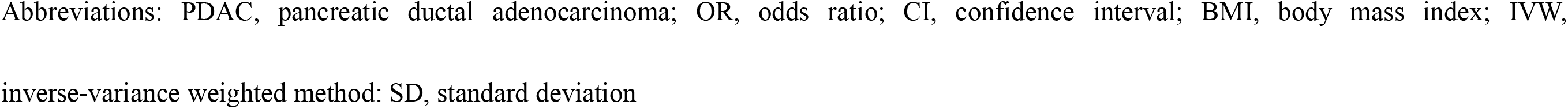
Comprehensive Mendelian randomization estimates of the association between pancreas fat and PDAC

| No. of Genetic Variants | Analysis | Association of pancreas fat with PDAC |  |  |  |
| --- | --- | --- | --- | --- | --- |
|  |  | OR (95% CI)<sup>c</sup> | P value | P for pleiotropy<sup>d</sup> | P for distortion<sup>e</sup> |
| 8 | IVW | 2.46 (1.38, 4.40) | 0.002 |  |  |
| 8 | Weighted median | 1.79 (1.13, 2.83) | 0.013 |  |  |
| 8 | MR-Egger | 4.56 (0.14, 144.9) | 0.39 | 0.72 |  |
| 6 (exclusion of outlier variants<sup>a</sup>) | MR-PRESSO | 2.29 (1.61, 3.26) | < 0.001 |  | 0.57 |
| 5 (exclusion of variants nominally linked to BMI<sup>b</sup>) | IVW | 3.79 (1.66, 8.65) | 0.002 |  |  |

<sup>a</sup>Two outlier genetic variants (chr9:136138765 and rs10422861) were detected in MR-PRESSO.
<sup>b</sup>Although none of the eight genetic variants were associated with BMI at genome-wide significance, we further conducted a sensitivity analysis excluding three genetic variants with a nominal BMI association.
<sup>c</sup>OR (95%CI) of PDAC per 1 SD increase in genetically determined pancreas fat levels (i.e., per 7.9% increase in pancreas fat fraction).
<sup>d</sup>P for pleiotropy was obtained from P value of MR-Egger intercept. Less than 0.05 indicates a possible pleiotropic effect.
<sup>e</sup>P for distortion was obtained from MR-PRESSO. Less than 0.05 indicates a difference between estimates before and after exclusion of outlier genetic variants.
Abbreviations: PDAC, pancreatic ductal adenocarcinoma; OR, odds ratio; CI, confidence interval; BMI, body mass index; IVW, inverse-variance weighted method; SD, standard deviation

## Discussion

Our Mendelian randomization study demonstrates a causal relationship between pancreas fat accumulation and PDAC. Consequently, pancreas fat represents a novel pathogenic contributor of this disease. This is likely independent of general adiposity, as suggested by our sensitivity analysis that excluded genetic variants nominally linked to BMI.

While our study is the first to provide genetic evidence for causality using Mendelian randomization, it is well in line with earlier conventional epidemiologic work on the topic ^16, 37, 38^. These studies reported that pancreas fat is more frequently found in patients with precancerous lesions (i.e., intraepithelial neoplasia) or PDAC, and can even be a predictor for PDAC ^16, 37, 38^. Similar to our current findings, these prior studies also observed that the relationship between pancreas fat and PDAC is independent of overall body fat (i.e., BMI) ^16, 37^. Thus, our data along with previous work support the idea that adipocytes within the pancreas may have unique features that differ from adipocytes in other locations, as suggested by basic science ^5, 39^.

One possible mechanism linking pancreas fat and PDAC is through the enhanced production of cytokines and adipokines from adipocytes residing within the pancreas ^5^. By stimulating inflammation, suppressing apoptosis, and promoting cell proliferation and migration, these cytokines and adipokines can contribute to cancer development or progression ^7, 40^. In fact, a prospective cohort study using ultrasonography reported that pancreas fat is a risk factor for future subclinical chronic pancreatitis ^41^, supporting the hypothesis that pancreas fat can contribute to chronic low-grade inflammation of the pancreas, a well-established driver of PDAC ^7, 13^. Another potential mechanism linking pancreas fat and PDAC is one mediated through diabetes mellitus. Excess pancreas fat can impair insulin secretion from beta cells in pancreatic islets and may lead to the development of diabetes mellitus ^4, 12, 13^, another long-known risk factor for PDAC that has also been causally associated with PDAC in a past Mendelian randomization study ^42^. Furthermore, accumulating evidence suggests that adipocytes in the pancreas exhibit marked heterogeneity in their secreted cytokines and adipokines between individuals, in part depending on systemic metabolism, as well as regulatory circulating factors from other organs ^4^. Hence, there appears to be a complex relationship between pancreas fat, inflammation, diabetes mellitus, and PDAC that needs to be disentangled by further mechanistic research.

Our current findings can have major clinical implications as excess pancreas fat accumulation is a reversible condition ^43^. A substantial weight loss through bariatric surgery or hypocaloric diet results in the reduction of pancreas fat ^13^, and an innovative diet intervention also shows a specific effect on this fat compartment ^44^. Promisingly, emerging pharmacological therapies for weight loss have been shown to significantly reduce fat mass ^45, 46^, suggesting that they may also have the potential to lower pancreas fat. Further studies are needed to clarify if and to what extent a reduction of pancreas fat ultimately translates into decreased PDAC incidence.

One major strength of this study is the use of Mendelian randomization, which is a robust approach less susceptible to reverse causation and confounding compared to conventional observational studies. Secondly, we utilized data from the largest GWAS on pancreas fat and PDAC to date. Third, we also incorporated a third data source (i.e., GIANT) to address potential pleotropic associations with BMI, a potential confounder, which could have biased our Mendelian randomization findings. Lastly, we used pancreas fat data measured with MRI, which has been well validated against histologic pancreas fat measurements and the most sensitive non-invasive modality for detection of pancreas fat ^10^.

There are some limitations to our study. The unknown exact mechanism linking the genetic variants, pancreas fat, and PDAC could theoretically involve pleiotropic effects that may violate assumptions of Mendelian randomization. To minimize the influence of this, we confirmed the robustness of our results through several sensitivity analyses including leave-one-out analyses, pleiotropy-robust statistical methods, and the analysis excluding genetic variants nominally linked to BMI. Another limitation is the restriction to individuals of European ancestry. Future studies should expand this analysis to non-European populations, as well as incorporate genetic variants for pancreas fat identified in other racial/ethnic groups ^47^. Lastly, our Mendelian randomization analysis detected the association of genetic predisposition to lifetime accumulation of pancreas fat with PDAC risk; however, our approach cannot clarify the impact of shorter-term changes in pancreas fat.

In conclusion, excess pancreas fat accumulation is a novel risk factor that causally contributes to PDAC, likely independent of overall body fat. Mechanisms linking pancreas fat to PDAC may include inflammatory and cancer-promoting signals from the local adipocytes. Pancreas fat may serve as an easily measurable and non-invasive biomarker of PDAC risk and may be even of greater utility in individuals with already elevated risk due to other reasons, such as chronic pancreatitis, adult-onset diabetes, inheritance of predisposing mutations, or family history ^14^. More importantly, our work further raises the possibility that reduction of pancreas fat could lower the incidence of PDAC.

## Data Availability

The pancreas fat genetic dataset is available from a previous study (reference 21): https://cdn.elifesciences.org/articles/65554/elife-65554-supp1-v1.xlsx. The pancreatic cancer genetic datasets used for the association analyses described in this manuscript were obtained from dbGaP at https://www.ncbi.nlm.nih.gov/gap/ through dbGaP accession phs000206.v5.p3 and phs000648.v1.p1. The BMI genetic dataset used for this study was obtained from a previous study (reference 25): https://zenodo.org/record/1251813#.Y9n8YnbP1D8.

## Acknowledgements

The authors thank Dr. Hua Zhong at University of Hawaii Cancer Center for her help for this study. We thank to participants and investigators of the UK Biobank and GIANT consortium for making GWAS summary statistics publicly available. The authors also would like to thank all the individuals for their participation in the parent studies and all the researchers, clinicians, technicians and administrative staff for their contribution to the studies. The PanScan study was funded in whole or in part with federal funds from the National Cancer Institute (NCI), US National Institutes of Health (NIH) under contract number HHSN261200800001E. Additional support was received from NIH/NCI K07 CA140790, the American Society of Clinical Oncology Conquer Cancer Foundation, the Howard Hughes Medical Institute, the Lustgarten Foundation, the Robert T. and Judith B. Hale Fund for Pancreatic Cancer Research and Promises for Purple. A full list of acknowledgments for each participating study is provided in the Supplementary Note of the manuscript with PubMed ID: 25086665. For the PanC4 GWAS study, the patients and controls were derived from the following PanC4 studies: Johns Hopkins National Familial Pancreas Tumor Registry, Mayo Clinic Biospecimen Resource for Pancreas Research, Ontario Pancreas Cancer Study (OPCS), Yale University, MD Anderson Case Control Study, Queensland Pancreatic Cancer Study, University of California San Francisco Molecular Epidemiology of Pancreatic Cancer Study, International Agency of Cancer Research and Memorial Sloan Kettering Cancer Center. This work is supported by NCI R01CA154823. Genotyping services were provided by the Center for Inherited Disease Research (CIDR). CIDR is fully funded through a federal contract from the National Institutes of Health to the Johns Hopkins University, contract number HHSN2682011000111. The content is solely the responsibility of the authors and does not necessarily represent the official views of the National Institutes of Health.

## Data availability

The pancreas fat genetic dataset is available from a previous study^21^: https://cdn.elifesciences.org/articles/65554/elife-65554-supp1-v1.xlsx. The pancreatic cancer genetic datasets used for the association analyses described in this manuscript were obtained from dbGaP at https://www.ncbi.nlm.nih.gov/gap/ through dbGaP accession phs000206.v5.p3 and phs000648.v1.p1. The BMI genetic dataset used for this study was obtained from a previous study^25^: https://zenodo.org/record/1251813#.Y9n8YnbP1D8.

## Abbreviations

(BMI): body mass index
(CT): computed tomography
(CI): confidence intervals
(GWAS): genome-wide association studies
(GIANT): Genetic Investigation of Anthropometric Traits
(InSIDE): instrument strength independent of direct effect
(IVW): inverse-variance weighted
(MRI): magnetic resonance imaging
(MAF): minor allele frequency
(OR): odds ratio
(PDAC): pancreatic ductal adenocarcinoma
(PanC4): Pancreatic Cancer Case-Control Consortium
(PanScan): Pancreatic Cancer Cohort Consortium
(SD): standard deviation

---

**Supplementary Figure 1.**
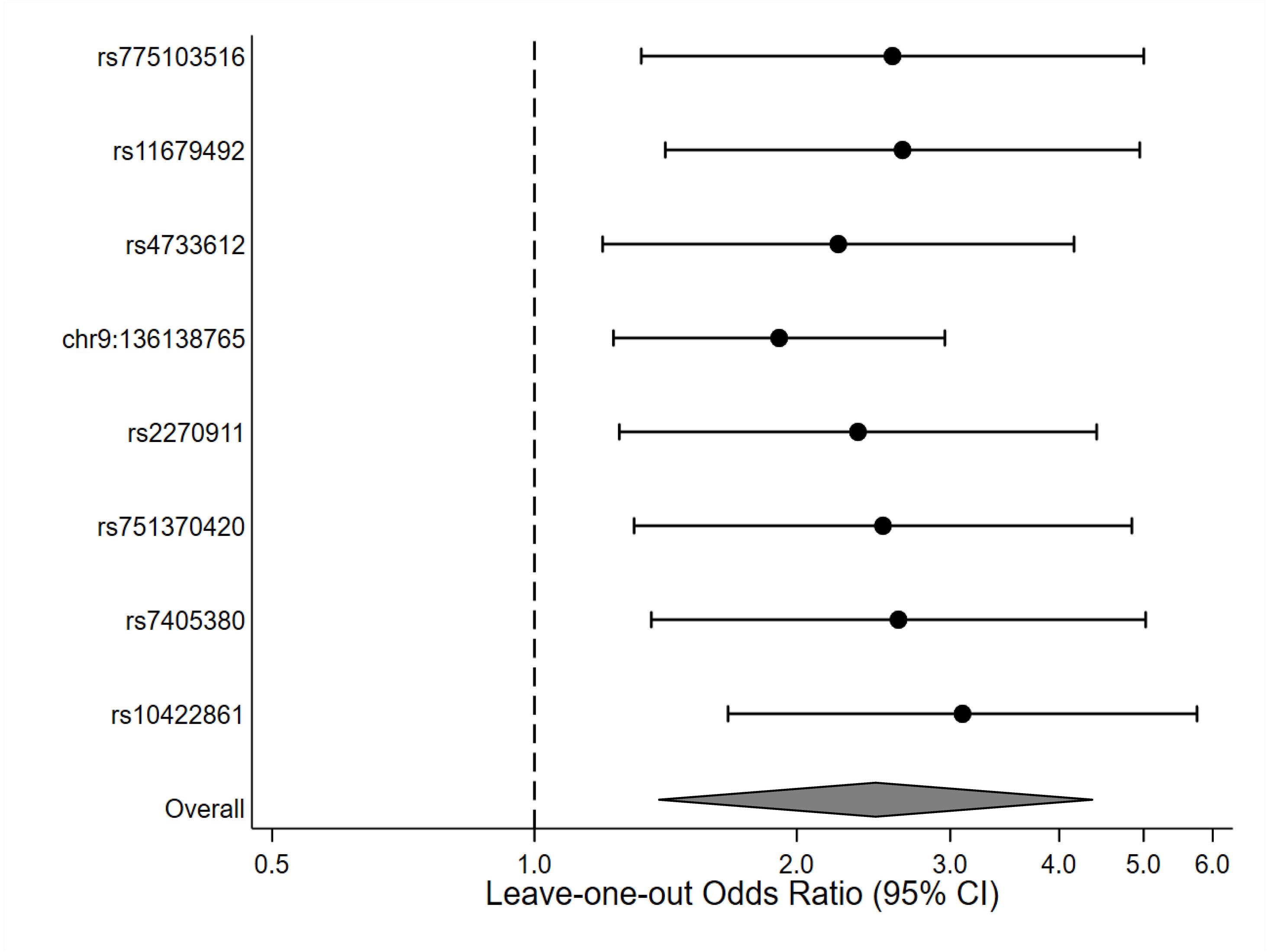
Leave-one-out Mendelian randomization estimates of the association between pancreas fat and PDAC. For leave-one-out analysis, each genetic variant was excluded, and we used random-effects inverse-variance weighted method to estimate odds ratio (95% CI) for the association between genetically determined pancreas fat and PDAC with the remaining genetic variants. As an example, for the top row, rs775103516 was excluded, and odds ratio (95% CI) was estimated using the remaining 7 genetic variants. Proxy single-nucleotide polymorphisms were used for rs775103516 (rs113170275), chr9:136138765 (rs495828), rs751370420 (rs7307879), and rs7405380 (rs12444726). Random-effects inverse-variance weighted method was used to estimate odds ratios (95% CI) of PDAC per 1 SD increase in genetically determined pancreas fat levels (i.e., per 7.9% increase in pancreas fat fraction). Data markers indicate the odds ratio for the association of genetically determined pancreas fat with PDAC. Error bars indicate 95% CIs. Abbreviations: PDAC, pancreatic ductal adenocarcinoma; CI, confidence interval; SD, standard deviation

